# Usual-Paced 400m Long Distance Corridor Walk Estimates Cardiorespiratory Fitness among Older Adults: The Study of Muscle, Mobility and Aging (SOMMA)

**DOI:** 10.1101/2023.11.05.23298103

**Authors:** Reagan E. Moffit, Peggy M. Cawthon, Barbara J. Nicklas, Bret H. Goodpaster, Paul M. Coen, Daniel E. Forman, Steven R. Cummings, Anne B. Newman, Nancy W. Glynn

## Abstract

**Background:** Cardiopulmonary exercise testing (CPET), the gold-standard method to quantify cardiorespiratory fitness (CRF), is not always feasible due to cost, access, and burden. The usual-paced 400m long-distance corridor walk (LDCW), a measure of mobility among older adults, may provide an alternate method to assess CRF among populations unable to complete maximal intensity testing. The purpose of this study was to develop and validate an estimating equation to estimate VO_2_peak from average 400m walking speed (WS) among participants in the Study of Muscle, Mobility and Aging (SOMMA).

**Methods:** At baseline, participants (N=820, 76.2±4.9 years, 58% Women, 86% Non-Hispanic White) completed a 400m LDCW (400m WS=400m/completion time in seconds) and symptom-limited maximal CPET (Modified Balke Protocol). VO_2_peak (mL/kg/min) was considered the highest 30-second average oxygen consumption during CPET. Other covariates included: age, sex, race, physical activity (7-day wrist-worn accelerometer), physical function (Short Physical Performance Battery, range 0-12), perceived physical fatigability (Pittsburgh Fatigability Scale, range 0-50), and Borg Rating of Perceived Exertion (RPE, range 6-20) at completion of the 400m LDCW. Stepwise linear regression was used. Internal validation was completed using data-splitting method (70%; 30%).

**Results:** Mean VO_2_peak was 20.2±4.8 mL/kg/min and mean 400m WS was 1.06±0.2 m/s. Each 0.05 m/s increment in 400m WS was associated with a 0.40 mL/kg/min higher VO_2_peak after adjustment for covariates. An estimating equation including 400m WS, age, sex, race, and RPE was developed. Internal validation showed low overall bias (-0.26) and strong correlation (r = 0.71) between predicted and measured VO_2_peak values. Bland-Altman plot and regression analyses indicated predicted VO_2_peak was an acceptable alternative, despite mean underestimation of 4.53 mL/kg/min among those with CPET VO_2_peak ≥25 mL/kg/min.

**Conclusions:** Usual-paced 400m LDCW strongly correlates with direct measures of cardiorespiratory fitness during CPET in older adults with lower fitness and can be used to test both fitness and function.

## INTRODUCTION

Cardiorespiratory fitness (CRF), the capacity of the cardiovascular and respiratory systems to supply oxygen to skeletal muscles for sustained activity, decreases with age, with accelerated declines occurring among those >70 years.^1–4^ Lower CRF is associated with higher risk of mortality and morbidity, namely cardiovascular disease, cancer, metabolic syndrome, and depression.^5–8^ Among older adults, understanding functional fitness is of interest as this may directly impact their activity levels, impacting clinical outcomes such as physical performance.

Peak oxygen uptake (VO_2_) is commonly used clinically and in research as an indicator of CRF. Cardio-pulmonary exercise testing (CPET), objective assessment of VO_2_ during progressive intensity exercise, is considered the gold-standard method of obtaining CRF.^9^ However, CPET testing is not always feasible in large-scale studies of older adults, due to rigorous staff training, expense, access, and participant burden. Because of these limitations, other methods are used as alternatives to CPET, including walking tests^10^ and non-exercise prediction equations.^11^ One such walking test developed for use among older adults is the validated fast-paced 400m long-distance corridor walk (LDCW), making it a good proxy for CRF.^10^

However, usual-paced 400m LDCW, a physical performance mobility measure and gold-standard method for identifying mobility disability (inability to complete 400m walk in <15 minutes)^12,13^ is often used in clinical trials and observational studies of older adults instead of a fast-paced 400m LDCW. This is primarily due to the impracticality of including both measures in clinic visits, as well as for safety concerns among older adults with higher levels of fatigue and frailty.^14^ Previous work has shown that fast-paced and usual-paced 400m LDCW completion times are comparable, especially among older and lower functioning adults as these groups may be working closer to their VO_2_peak while walking at their usual pace.^15^ Due to the importance of measuring CRF among older adults, it is key to assess whether a usual-paced 400m LDCW can estimate VO_2_peak.

Previously, we determined that faster usual-paced 400m LDCW completion was directly associated with higher VO_2_peak in a small, healthy population of older adults.^16^ However, limited sample size prohibited our ability to generalize our findings across a wider range of function and fitness. Therefore, we assessed whether a usual-paced 400m LDCW was associated with VO_2_peak among participants in the Study of Muscle, Mobility and Aging (SOMMA). We also developed and validated an estimating equation for VO_2_peak values from average 400m LDCW walking speed (400m WS). We hypothesized that there would be a strong positive association between 400m WS and VO_2_peak. We also compared equation performance by sex, physical function, and perceived physical fatigability.

## METHODS

### Study Sample

SOMMA (https://www.sommastudy.com) participants were recruited from two sites (University of Pittsburgh, Wake Forest University School of Medicine). Baseline eligibility included aged ≥70 years, body mass index (BMI) ≤40kg/m^2^, willing to undergo muscle tissue biopsy and magnetic resonance spectroscopy scan, and able to walk 400m in <15 minutes. Exclusion included self-reported inability to walk one-quarter mile or climb a flight of stairs, medical contraindication to biopsy or MR, individuals on chronic anticoagulation therapy, and those with active malignancies or advanced chronic disease (e.g. heart failure, dementia). A detailed study design paper is published elsewhere.^17^ Participants gave written informed consent, and the WIRB-Copernicus Group (WCG) Institutional Review Board (WCGIRB, study number 20180764) approved SOMMA as a single IRB.

### Usual-Paced 400m Long Distance Corridor Walk (LDCW)

Participants were instructed to “walk your usual pace” for 10 laps around a 40meter course (20 meters marked space with cones on either side) in a long corridor, without assistive devices other than a straight cane (n=22). Standard encouragement was provided throughout the LDCW. Participants were allowed to stop and rest standing in place for up to one minute during the test (n=11). Average 400m WS (m/s) was calculated as 400 (m) divided by completion time (s).

### Cardiorespiratory Fitness (VO_2_peak)

CRF was measured by CPET using a modified symptom-limited Balke protocol in which speed and grade increased incrementally.^18^ O_2_ consumption and CO_2_ exhalation were measured breath-by-breath using a Hans Rudolph face mask and cardiopulmonary metabolic cart (Medgraphics Ultima Series, Medgraphics Corporation, St. Paul, MN). Participants were verbally encouraged to reach a respiratory exchange ratio (RER; breath-by-breath measured VCO_2_ to VO_2_ ratio) of ≥1.05 and a Rating of Perceived Exertion (RPE)≥17 before test termination.^19^ Test was terminated in the event of significant symptoms (e.g. moderate-severe chest pain, increasing dizziness) or significant arrhythmia or ischemic changes on ECG. Highest 30 second average oxygen consumption (VO_2_peak) sustained during the CPET test was identified by the BREEZESUITE software and confirmed by the clinical exercise physiologist. VO_2_peak was normalized to weight (mL/kg/min). Adjudication took place if participants met these criteria: VO_2_peak mL/kg/min <12 ml/kg/min, maximum RPE <15 and maximum RER<1.05 (up to the start of recovery), or maximum heart rate <100 bpm (n=47; n=20 deemed valid).

### Covariates

Age and sex were self-reported. Participants also reported race/ethnicity. Physical function was assessed using the Short Physical Performance Battery (SPPB, range 0-12); worse=SPPB<10 vs better=SPPB≥10.^20^ Moderate vigorous physical activity (MVPA, minutes/week) was assessed for 7 days using a wrist-worn accelerometer (ActiGraph GT9X, valid wear=≥3 days of ≥10 hours).^21^ Perceived physical fatigability was assessed using the validated 10-item Pittsburgh Fatigability Scale (PFS, range 0-50); more severe=PFS≥15 vs less severe=PFS<15.^22,23^ Perceived exertion at the end of the 400m LDCW was measured using Borg RPE (range 6-20, higher=more exertion).

### Statistical Analysis

Descriptive statistics (mean, SD, frequencies) and Pearson correlations were calculated. We used linear regression to examine the association between 400m WS and VO_2_peak; final model was adjusted for age, sex, race (white vs non-white), MVPA, SPPB, PFS, and site (n=763, removed participants with missing covariate data). To develop and validate an estimating equation for VO_2_peak, participants with complete VO_2_peak and 400m WS (N=820) were randomly split into training (70%, n=574) and validation (30%, n=246) subsets. Forward selection stepwise regression was used to develop the final model in the training subset for estimating VO_2_peak from 400m WS. Only variables intrinsic to the usual-paced 400m LDCW were considered, plus commonly assessed demographics and anthropometric measures. Variables were initially selected after independently being associated with VO_2_peak (p<0.05) and then individually added to the equation and retained if p<0.05 and positively impacted the coefficient of determination (R^2^). We assessed the validity of predicted VO_2_ peak in the validation subset using Spearman correlation, bias (sum[predicted – measured]/n), Bland-Altman Plots, and regression analyses (good validity if intercept=0 and β=1). A t-test compared measured differences between those with CRF <25 mL/kg/min and ≥25 mL/kg/min. Performance was examined in the full sample (N=820) stratified by sex, SPPB, and PFS using Spearman correlation. Analyses were performed in R (version 4.2.3).

## RESULTS

A total of 820 SOMMA participants had valid VO_2_peak and 400m WS data at baseline (Supplemental Figure S1). On average, participants (76.2±4.9 years, 86% White, 58% Women, 27% lower functioning, SPPB<10) had a mean VO_2_peak of 20.2±4.8 mL/kg/min and mean 400m WS of 1.06±0.2 m/s (Table 1). Faster 400m WS and higher VO_2_peak were moderate-to-strongly correlated (r=0.55, p<0.01). A 0.05 m/s faster 400m WS was associated with a 0.78 mL/kg/min higher VO_2_peak, explaining 30.7% of the variance; association was attenuated after full adjustment (β=0.40 mL/kg/min), (Table 2). Full adjustment improved model fit (R^2^=0.54).

**Table 1.** Descriptive characteristics of Study of Muscle, Mobility and Aging (SOMMA) participants.

|  | **Overall (N=820)** |
| --- | --- |
|  | **Mean ± standard deviation or n (%)** |
| Age, years | 76.2±4.90 |
| Sex, Women | 478 (58) |
| Race, White | 709 (86) |
| Body mass index, kg/m<sup>2</sup> | 27.5±4.5 |
| Hypertension, SBP ≥140 mmHg | 226 (28) |
| Heart Disease* | 56 (6.9) |
| Lung Disease* | 104 (13) |
| Diabetes* | 118 (14) |
| Arthritis* | 458 (56) |
| Stroke* | 20 (2.4) |
| Myocardial Infarction* | 21 (2.6) |
| Peripheral Vascular Disease* | 6 (0.7) |
| Chronic Renal Disease or Renal Failure* | 30 (3.7) |
| No Problems Performing Usual Activities | 746 (92) |
| I Have No Problems with Self-Care | 811 (99) |
| I Have Some Problems Walking About | 106 (13) |
| MVPA, min/week | 188.5±84.9 |
| Average 400m Walking Speed, m/s | 1.06±0.2 |
| Time to Complete 400m LDCW, sec | 388.8±69.7 |
| Borg RPE at End of 400m LDCW, 6-20 | 9.7±2.2 |
| VO<sub>2</sub>peak, mL/kg/min | 20.2±4.8 |
| PFS Physical score, 0-50 | 15.5±8.6 |
| SPPB, 0-12 | 10.3±1.7 |

MVPA = accelerometry measured moderate-vigorous physical activity; Borg RPE = rating of perceived exertion; PFS Physical Score = Pittsburgh Fatigability Scale Physical Sub-Score; SPPB = Short Physical Performance Battery \*Self-report of a physician diagnosis

**Table 2.**
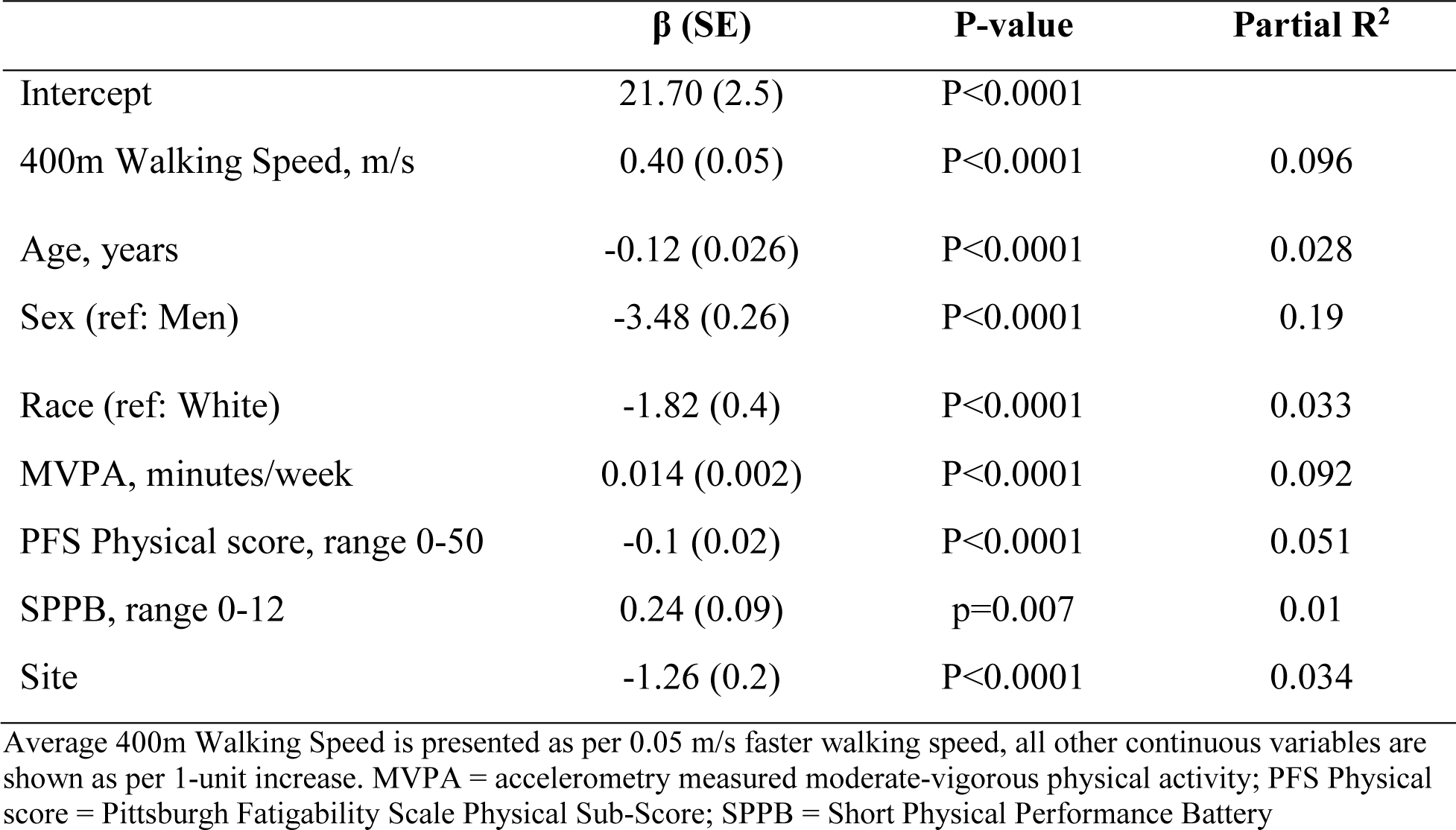
Fully adjusted model of the associations of VO_2_peak on average walking speed during usual-paced 400m LDCW among Study of Muscle, Mobility and Aging (SOMMA) participants (N=763)

### Development and Validation of Estimating Equation

The final estimating equation explaining 46.3% of the variance in measured VO_2_peak:

> *Predicted VO_2_peak = -19.40 + (55.34*400m WS) - (0.42*Ending RPE) + (0.44*Age) - (2.68*Sex) - (2.31*Race) - (0.58*400m WS*Age))*

The estimating equation performed the same or slightly better in the validation set (R^2^=0.49, r=0.71) compared to the training set (R^2^=0.46, r=0.68). Overall, predicted VO_2_peak bias was close to 0 at -0.26, indicating low group-wide bias. Regression analysis showed low potential for systematic bias (β = 0.97, p<0.001; Intercept = 0.95, p=0.47), although a test for heteroskedasticity was significant (p<0.05). The Bland-Altman plot showed a slightly negative sloping pattern and few points sitting outside the 95% CI (Figure 1). Those with measured VO_2_peak values ≥25 mL/kg/min had larger mean difference between measured and predicted VO_2_peak compared to those with measured VO_2_peak <25 mL/kg/min (-4.53 vs 0.62 mL/kg/min, p<0.0001), supporting underestimation of VO_2_peak by our estimating equation among those with higher CRF. To improve model fit for those with higher CRF, we used change-point analysis to identify an inflection-point between VO_2_peak and 400m WS; however, no clear inflection-point was identified (Figure S2).

**Figure 1.**
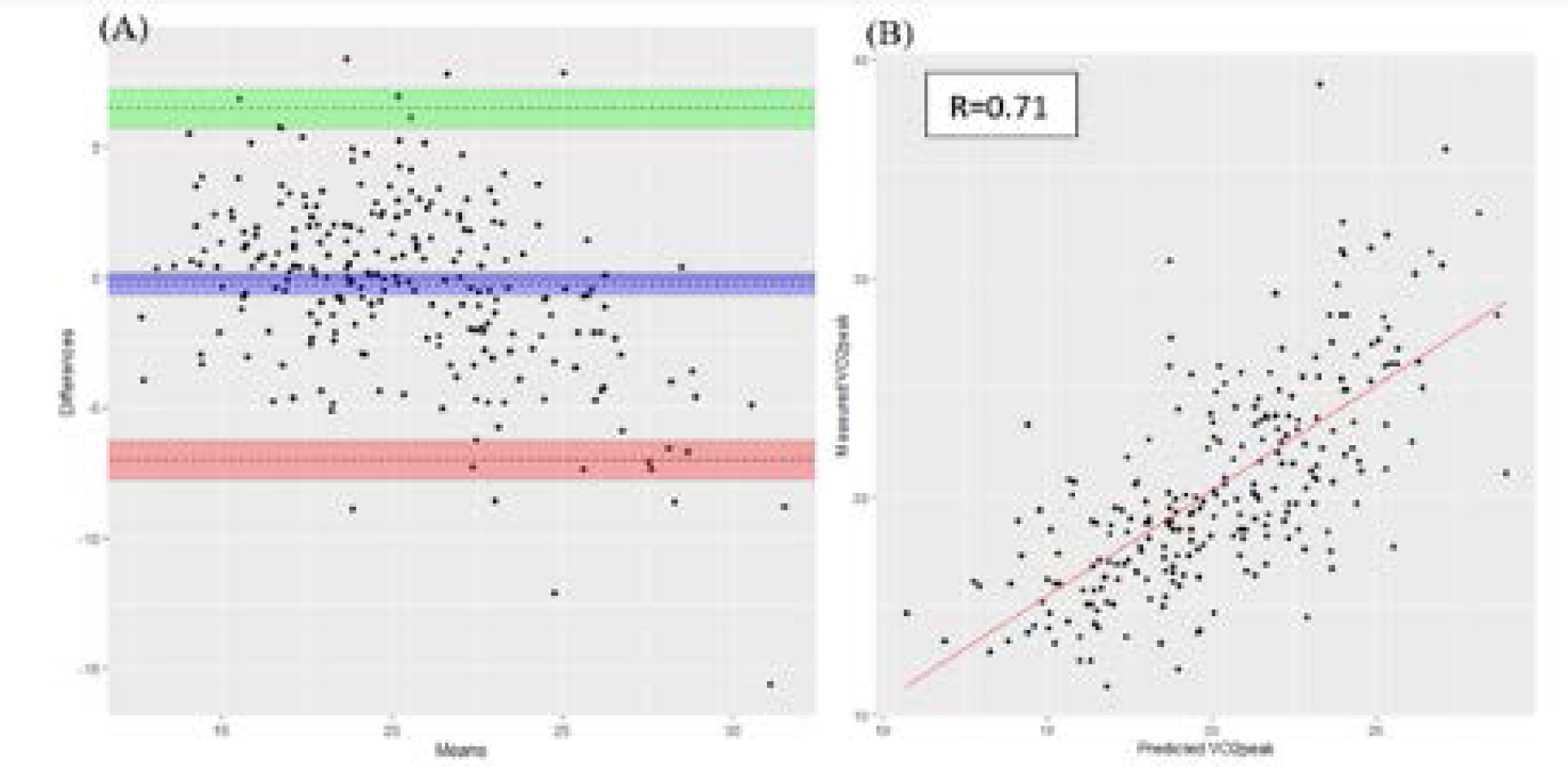
Visualizations of the internal validation measures of the estimating equation developed in the Study of Muscle, Mobility and Aging (SOMMA) Figure A depicts the Bland-Altman plot of measurement agreement among the entire sample (N=820); the x-axis is the mean value between measured and predicted VO_2_ peak values, and the y-axis is the difference between measured and predicted VO_2_ peak values. The horizontal line at 0 represents perfect agreement, whereas the dotted lines represent the mean measurement agreement (center dotted line and blue shaded area) and 95% Confidence Intervals from the mean measurement agreement, respectively (green and red shaded areas). Figure B visually represents the regression analyses performed for internal validation. The red solid line indicates the fitted lines for the analyses. R=correlation coefficient. Both plots were generated in the validation subset.

In the overall sample, the estimating equation performed similarly by sex (women: r=0.63; men: r=0.61), physical function (SPPB≥10: r=0.64; SPPB<10:r=0.62), and perceived physical fatigability (PFS<15: r=0.66; PFS≥15:r=0.61).

## DISCUSSION

Within a sample of community-dwelling older adults, 400m WS was associated with CRF, even after covariate adjustment. Our results suggest that the usual-paced 400m LDCW entails a physiologic workload that is sufficient to assess CRF in older adults across a wide range of function and fitness. We also successfully developed and validated an estimating equation to obtain VO_2_peak from the usual-paced 400m LDCW.

We previously found that usual-paced 400m LDCW completion time was correlated with (r=-0.64) and explained 41% of the total 57% variance in VO_2_peak.^16^ Our model in SOMMA explained slightly less variance, even after adjustment for more potential confounding variables. When instructed to walk at a usual-pace, some participants may walk at a speed that puts them below a certain intensity threshold to avoid physiological responses to exercise. For example, patients with heart failure choose walking speeds resulting in lower ventilation cost.^24^ Conversely, some older adults may walk at a speed that is tapping into their energy reserve due to older age or poor physical function.^25,26^ The large range of 400m WS (0.46-1.60 m/s) in this cohort may contribute to differential association between 400m WS and VO_2_peak, with subclinical diseases affecting the association among those with slower walking speeds.

We performed rigorous internal validation of the estimating equation. All measures indicated that our estimating equation is a valid alternative to traditional CPET, particularly among those with lower fitness. Correlation between predicted and measured VO_2_peak values was 0.71, compared to 0.87 observed in the fast-paced 400m LDCW.^10^ This was expected as the fast-paced 400m LDCW requires a higher walking intensity and thus greater reliance on CRF to complete. Additionally, the fast-paced 400m LDCW estimating equation included measures of physiologic effort and stride length, which provided additional context to the relative effort of the task, thus improving prediction of VO_2_peak. SOMMA did not collect measures of physiologic effort (e.g., heart rate) during the LDCW. We considered stride length in our estimating equation, but it was not retained as it was not statistically significant in the final model, likely due to 400m WS inherently accounting for stride length in the calculation of WS. We also purposefully did not consider any significant lifestyle variables (i.e., physical activity) as to not introduce extra assessments, cost, or time to derive estimated VO_2_peak values. Our estimating equation performed similarly to other walk-based submaximal tests, such as the 1-mile (r=0.62) or 15-minute walk test (r=0.42).^27^ Using a usual-paced 400m LDCW test may be preferable in older, less fit adults free of mobility disability because the distance to complete is shorter and pace is not as strenuous.

As shown by the negative sloping trend in the Bland-Altman plot and the t-test between measured differences, our estimating equation underestimated VO_2_peak on average by 4.53 mL/kg/min among those with CRF ≥25mL/kg/min. Similar to the approach by Simonsick et al,^10^ attempts to improve model fit using a correction factor for 400m WS estimation of VO_2_peak did not significantly improve predicted VO_2_peak. Maintaining a higher VO_2_peak as one ages may be associated with better health status and greater physical activity level. In a usual-paced 400m LDCW, highly fit older adults are not working at a high enough intensity to be related to their VO_2_peak. Because the test protocol discouraged running, a ceiling effect is possible. While few older adults have high VO_2_peak,^28^ an understanding of the functional capacity of the target population is needed before applying our estimating equation in future studies. Studies that include those with higher fitness may want to directly measuring VO_2_peak or consider other submaximal tests, such as the fast-paced 400m LDCW.^10^

A strength of SOMMA was the ability to perform gold-standard maximal CPET testing in older adults using a rigorous protocol, enabling us to validate an estimating equation against the gold-standard in a cohort with a wide range of physical function. Further, the use of data-splitting allowed for internal validation of the estimating equation. The large analytic sample size (N=820) allowed for adequate power to be retained even after data-splitting. Though our estimating equation produced predicted VO_2_peak values that were strongly correlated with measured VO_2_peak values, the model explained 46% and 49% of the variance in the Training subset and Validation subsets, respectively. Including additional variables in the estimating equation could be considered to increase the model fit. SOMMA excluded participants with major mobility disability, limiting the generalizability to this group of older adults. SOMMA also had relatively small numbers of participants from underrepresented populations so whether our prediction equation performs similarly in different race/ethnic subgroups is not clear.

Our work extends the utility of a usual-paced 400m LDCW as a valid indicator of maximal CRF in older adults with lower fitness levels. Future epidemiologic studies that include less fit older adults may utilize the usual-paced 400m LDCW as both a test of fitness and function. Usual-paced 400m LDCW may be a better indicator of functional CRF as opposed to maximal CRF. Understanding functional CRF among older adults is particularly of interest as it is one of the most sensitive biomarkers of longevity and health. Consistent attainment of CRF may enhance approaches for therapeutics, decision making, and many other fundamental aspects of care. Future work warrants investigating the association of usual-paced 400m WS with elemental cellular determinants of CRF that underlie differences between vitality, functional decline, and disease.

## Data Availability

All data produced in the present study are available upon reasonable request to the authors.

https://sommaonline.ucsf.edu

## Conflict of Interest

The authors have no conflicts of interest to disclose.

## Author Contributions

Ms. Moffit and Dr. Glynn had full access to all of the data for this project and take responsibility for the integrity of the data and accuracy of the data analyses. Ms. Moffit drafted manuscript under Dr. Glynn’s supervision. All authors: interpretation of data, critical revision of manuscript for important intellectual content. All authors read and approved the submitted manuscript.

## Sponsor’s Role

none

## Supplemental Material for

**Figure S1.**
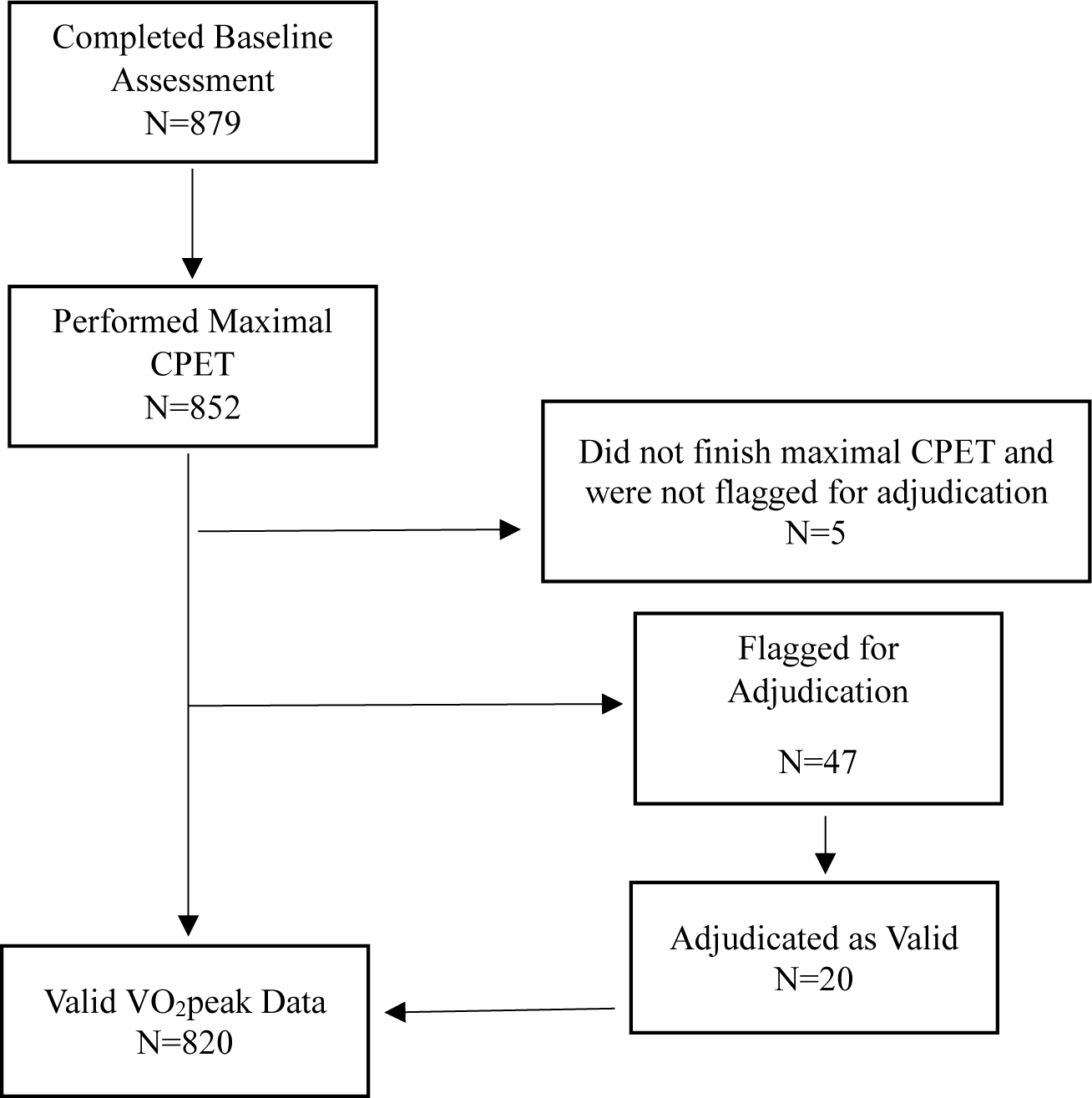
Sample Size of Current Analyses from the Study of Muscle, Mobility and Aging (SOMMA) The total sample size of SOMMA at baseline was 879 participants. Only participants with completed 400m Long-Distance Corridor Walk and valid VO2peak values were included in the current paper. No participants were unable to complete the usual-paced 400m LDCW. Of the 852 participants who attempted to complete the maximal Cardiopulmonary Exercise Test (CPET), 820 had valid VO2peak values. Adjudication occurred if VO_2_peak mL/kg/min <12 ml/kg/min, maximum RPE <15 and maximum RER<1.05 (up to the start of recovery), or maximum HR <100 bpm. Only one adverse event occurred during the maximal CPET (fall). No major injuries were sustained as a result of the fall.

**Figure S2.**
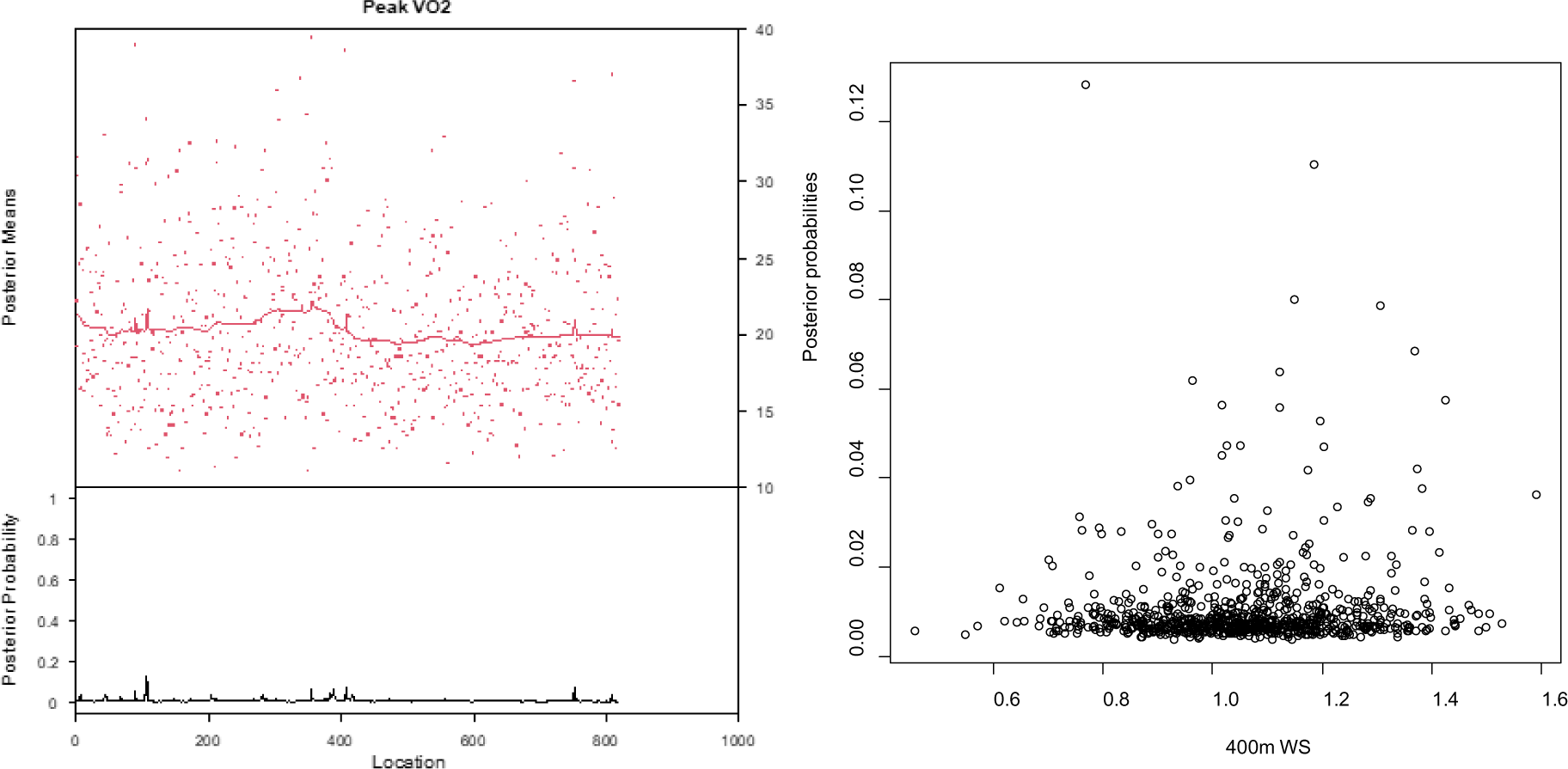
Results from Change-Point Analysis to Identify an Inflection Point between VO_2_peak and 400m Walk Speed in the Study of Muscle, Mobility and Aging (SOMMA) These figures depict results from the change-point analyses using the ‘bcp” R package. Figure A depicts the posterior probabilities and posterior means of VO2peak. Posterior probabilities can range from 0 to 1; a higher posterior probability indicates higher likelihood of change-point. Figure B is a scatterplot that shows the posterior probabilities by 400m WS. Higher posterior probability indicates higher likelihood of being a change-point. The highest posterior probability identified was 0.12 at 400m WS=0.76 m/s, indicating low likelihood of change-point. Including a correction factor where CF = 0.76 – 400m WS was not statistically significant in our estimating equation model.

## Notes

### Competing Interest Statement

The authors have declared no competing interest.

### Funding Statement

The Study of Muscle, Mobility and Aging is supported by funding from the National Institute on Aging (AG 059416). Study infrastructure support was funded in part by NIA Claude D. Pepper Older American Independence Centers at University of Pittsburgh (P30 AG024827) and Wake Forest University (P30 AG021332) and the Clinical and Translational Science Institutes, funded by the National Center for Advancing Translational Science, at Wake Forest University (UL1 0TR001420). Additionally, the Claude D. Pepper Older Americans Independence Center, Research Registry and Developmental Pilot Grant (NIH P30 AG024827), and the Intramural Research Program, National Institute on Aging supported N.W.G to develop the Pittsburgh Fatigability Scale.

### Author Declarations

Participants gave written informed consent, and the WIRB-Copernicus Group (WCG) Institutional Review Board (WCGIRB, study number 20180764) approved SOMMA as a single IRB.

